# Upper respiratory tract SARS-CoV-2 RNA loads in symptomatic and asymptomatic children and adults

**DOI:** 10.1101/2021.03.03.21252814

**Authors:** Rosa Costa, Felipe Bueno, Eliseo Albert, Ignacio Torres, Silvia Carbonell-Sahuquillo, Ana Barrés-Fernández, David Sánchez, Carmelo Padrón, Javier Colomina, María Isabel Lázaro Carreño, José Rafael Bretón-Martínez, Cecilia Martínez-Costa, David Navarro

## Abstract

**Objectives:** There is limited information comparing SARS-CoV-2 RNA load in the upper respiratory tract (URT) between children and adults, either presenting with COVID-19 or asymptomatic. Here we conducted a retrospective, single center study involving a large cohort of SARS-CoV-2 infected individuals to address this issue.

**Patients and Methods:** A total of 1,184 consecutive subjects (256 children and 928 adults) testing positive for SARS-COV-2 RNA in nasopharyngeal exudates (NP) were included, of whom 424 (121 children and 303 adults) had COVID-19 not requiring hospitalization and 760 (135 children and 625 adults) were asymptomatic close contacts of COVID-19 patients. SARS-CoV-2 RNA testing was carried out using the TaqPath COVID-19 Combo Kit (Thermo Fisher Scientific, MS, USA). The AMPLIRUN® TOTAL SARS-CoV-2 RNA Control (Vircell SA, Granada, Spain) was used for estimating SARS-CoV-2 RNA loads (in copies/mL).

**Results:** Median SARS-COV-2 RNA loads were comparable between adults and children with COVID-19 (7.14 log_10_ copies/ml vs. 6.98 log_10_ copies/ml; *P*=0.094). Median SARS-CoV-2 RNA load in asymptomatic children and adults was similar (6.20 log_10_ copies/ml vs. 6.48 log_10_ copies/ml; *P*=0.97). Children with COVID-19 symptoms displayed SARS-CoV-2 RNA loads comparable to their asymptomatic counterparts (*P*=0.61). Meanwhile in adults, median SARS-CoV-2 RNA load was significantly higher in symptomatic than in asymptomatic subjects (*P*=<0.001), yet comparable (*P*=0.61) when the analysis excluded patients sampled within 48 h after symptoms onset.

**Conclusions:** The data suggest that children may be drivers of SARS-CoV-2 transmission in the general population at the same level as adults.

## INTRODUCTION

An increasing body of evidence suggests that children are less susceptible to SARS-CoV-2 infection and tend to develop milder forms of COVID-19 than adults [1]. Nevertheless, whether children, either symptomatic or asymptomatic, play a major role in community transmission of SARS-CoV-2 compared to adults remains unclear [1]. There is a consistent direct correlation between magnitude of SARS-CoV-2 RNA load in the upper respiratory tract (URT) and probability of recovering live virus in cell culture, in both adults and children [2–5]; hence, viral load in URT may be used as a proxy for contagiousness. Supporting this assumption, transmission risk was recently shown to be strongly associated with initial SARS-CoV-2 RNA levels of index cases [6]. There is scarce information on how SARS-CoV-2 RNA load in UTR compares between children and adults [7–11], whether viral load in pediatric subjects differ across ages [9,10], and whether dissimilarities in the dynamics of SARS-CoV-2 shedding in URT exist between symptomatic and asymptomatic children [12–14]. Elucidation of these questions is critically important for designing effective public health policies to fight the pandemic. Here, to gain a further insight into these issues, we conducted a retrospective, single center study involving a substantial cohort of SARS-CoV-2 infected children and adults, either asymptomatic or symptomatic, non-hospitalized cases.

## METHODS

### Patients and specimens

A total of 1,184 consecutive subjects testing positive for SARS-COV-2 RNA in nasopharyngeal exudates (NP) between June 2020 and January 2021 were included. Participants were pediatric individuals (≤18 years; n=256; 21.6%), aged a median of 12 years (range, 0-18 years) or adults (>18 years; n=928; 78.3%), aged a median age of 37 years (range, 19-93 years). A total of 967 participants (154 children and 813 adults) were sampled at primary health centers belonging to the Health Department Clínico-Malvarrosa, Valencia (Spain), while 217 (102 children and 115 adults) were sampled at the Emergency Department of Hospital Clínico Universitario of Valencia. A total of 424 subjects (children, n=121; adults, n=303) presented with symptoms compatible with COVID-19, including one or more of the following: fever, dry cough, rhinorrhea, dyspnea, myalgia, fatigue, anosmia, ageusia, odynophagia, diarrhea, conjunctivitis, and cephalea, none requiring hospitalization. A total of 760 participants (135 children and 625 adults) were asymptomatic close contacts of COVID-19 patients, as previously defined [15]. NP specimen collection in the latter group was prescribed at the discretion of either the physician in charge of the index case or local health authorities, and was performed at a median of 7 days (range, 1-10 days) after diagnosis of the presumed index case. Initial SARS-CoV-2 RNA loads were used throughout the current study for comparison purposes. The current study was approved by the Research Ethics Committee of Hospital Clínico Universitario INCLIVA (March 2020).

### SARS-CoV-2 RNA testing

NPs were collected by trained nurses at sampling sites and were placed in 3 mL of Universal Transport Medium (Becton Dickinson, Sparks, MD, USA). RT-PCRs were carried out at the Microbiology Service of Hospital Clínico Universitario within 24 h of specimen collection. The TaqPath COVID-19 Combo Kit (Thermo Fisher Scientific, MS, USA), which targets SARS-CoV-2 ORF1ab, N and S genes, was used following RNA extraction carried out using the Applied Biosystems™ MagMAX™ Viral/Pathogen II Nucleic Acid Isolation Kits coupled with Thermo Scientific™ KingFisher Flex automated instrument. The AMPLIRUN® TOTAL SARS-CoV-2 RNA Control (Vircell SA, Granada, Spain) was used as the reference material for estimating SARS-CoV-2 RNA load (in copies/mL, taking RT-PCR CTs for the N gene) [16,17].

### RT-PCR β□glucuronidase RNA testing

We amplified the β□glucuronidase (GUSB) housekeeping gene to assess specimen cellularity in selected specimens following a previously published protocol [18]. In brief, RNA was extracted from NP using the DSP virus Pathogen Minikit on the QiaSymphony Robot instruments (Qiagen, Valencia, CA), reverse□transcribed to complementary DNA and subsequently amplified by using the HEQC one□step kit (Seqplexing, Valencia, Spain) in the LightCycler 480 Real□Time PCR System Version II (Roche Diagnostics, Pleasanton).

### Statistical methods

Differences between medians across groups were compared in a pairwise fashion using the non-parametric Mann–Whitney U-test, given that SARS-COV-2 RNA loads were non-normally distributed. Spearman’s rank test was used to test the association between age and SARS-CoV-2 RNA load. Two-sided exact *P*-values were reported. A *P*-value <0.05 was considered statistically significant. The analyses were performed using SPSS version 20.0 (SPSS, Chicago, IL, USA).

## RESULTS

### SARS-CoV-2 RNA load in pediatric and adult COVID-19 patients

We first compared initial SARS-CoV-2 RNA load in NP from symptomatic pediatric and adult patients. Specimen collection was carried out at a median of 2 days (range, 0-10 days) and a median of 3 days (range, 0-10 days), respectively, after symptoms onset. The data are shown in Fig. 1A. The range of estimated SARS-COV-2 RNA loads appeared comparable between children and adults; nevertheless, a trend towards a lower median viral RNA load was observed in children compared to adults (6.98 log_10_ copies/ml and 7.14 log_10_ copies/ml and), although the difference did not reach statistical significance (*P*=0.094).

**Figure 1.**
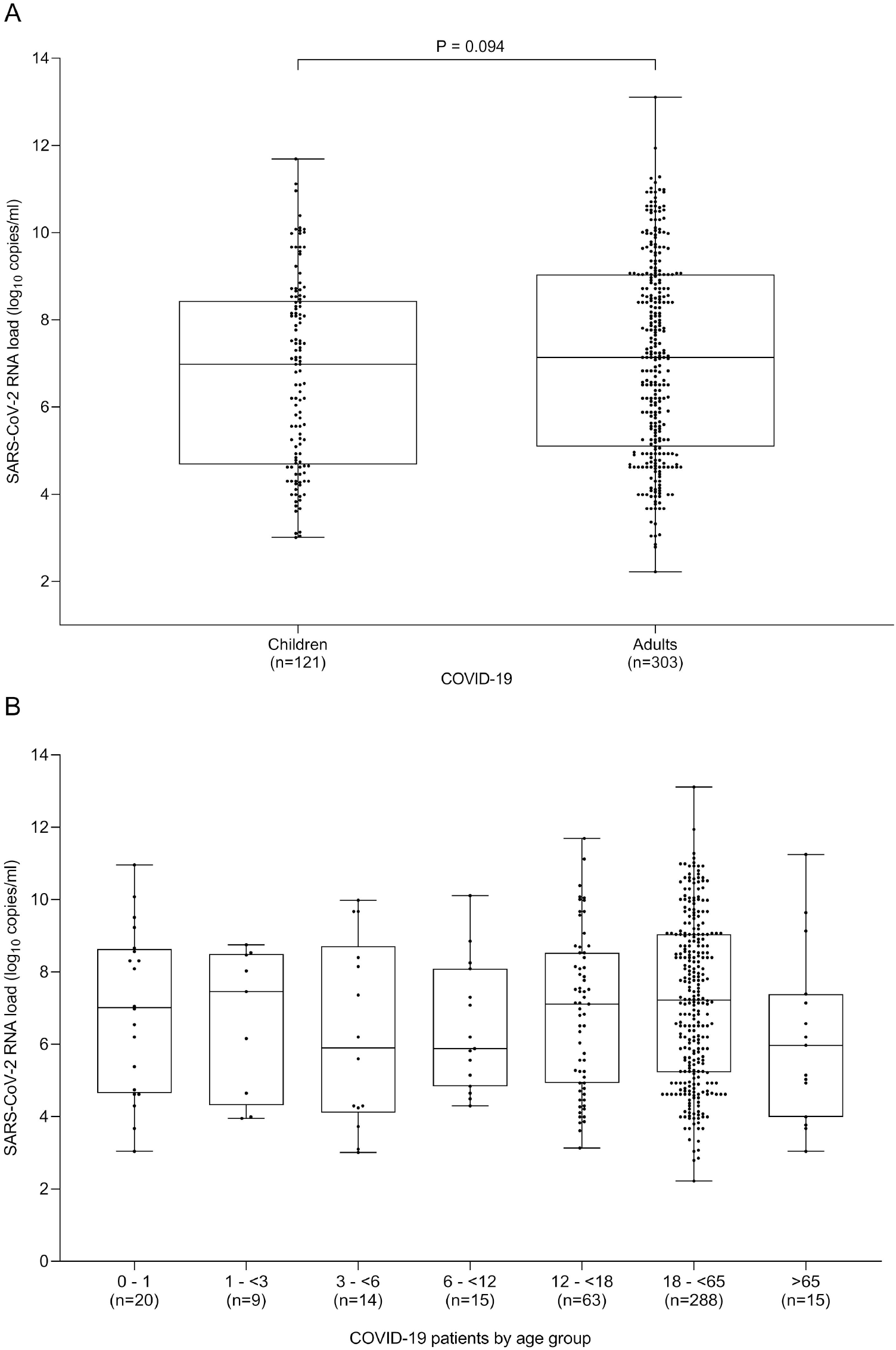
Overall estimated initial SARS-CoV-2 RNA loads in nasopharyngeal specimens from children and adults with COVID-19 (A) and those found across different pediatric and adult ages (B). Medians are indicated by midlines, the top and bottom edges of boxes represent the interquartile range (IQR). Whiskers indicate the upper and lower values. The number of patients in each group as well as *P* values for comparisons between groups (median SARS-CoV-2 RNA levels) are shown.

We next compared initial SARS-CoV-2 RNA load in children and adults by time of NP sampling after symptoms onset. Since SARS-CoV-2 RNA load peaks within the first 48 h after COVID-19 clinical presentation [18], we split each patient group into two subgroups (<3 days/≥3 days). As anticipated, SARS-CoV-2 RNA load was significantly higher in NP specimens collected within 48 h after onset of symptoms than in those obtained later on, in both children (median, 7.46 log_10_ copies/ml vs. 5.17 log_10_ copies/ml; *P*=<0.001) and adults (7.81 log_10_ copies/ml vs. 6.45 log_10_ copies/ml; *P*=0.002) (Fig. 2). Interestingly, SARS-CoV-2 RNA load measured within 48 h after symptoms onset was comparable (*P*=0.263) between children and adults, whereas those determined at later times (>48 h) were significantly lower in children (*P*=0.002).

**Figure 2.**
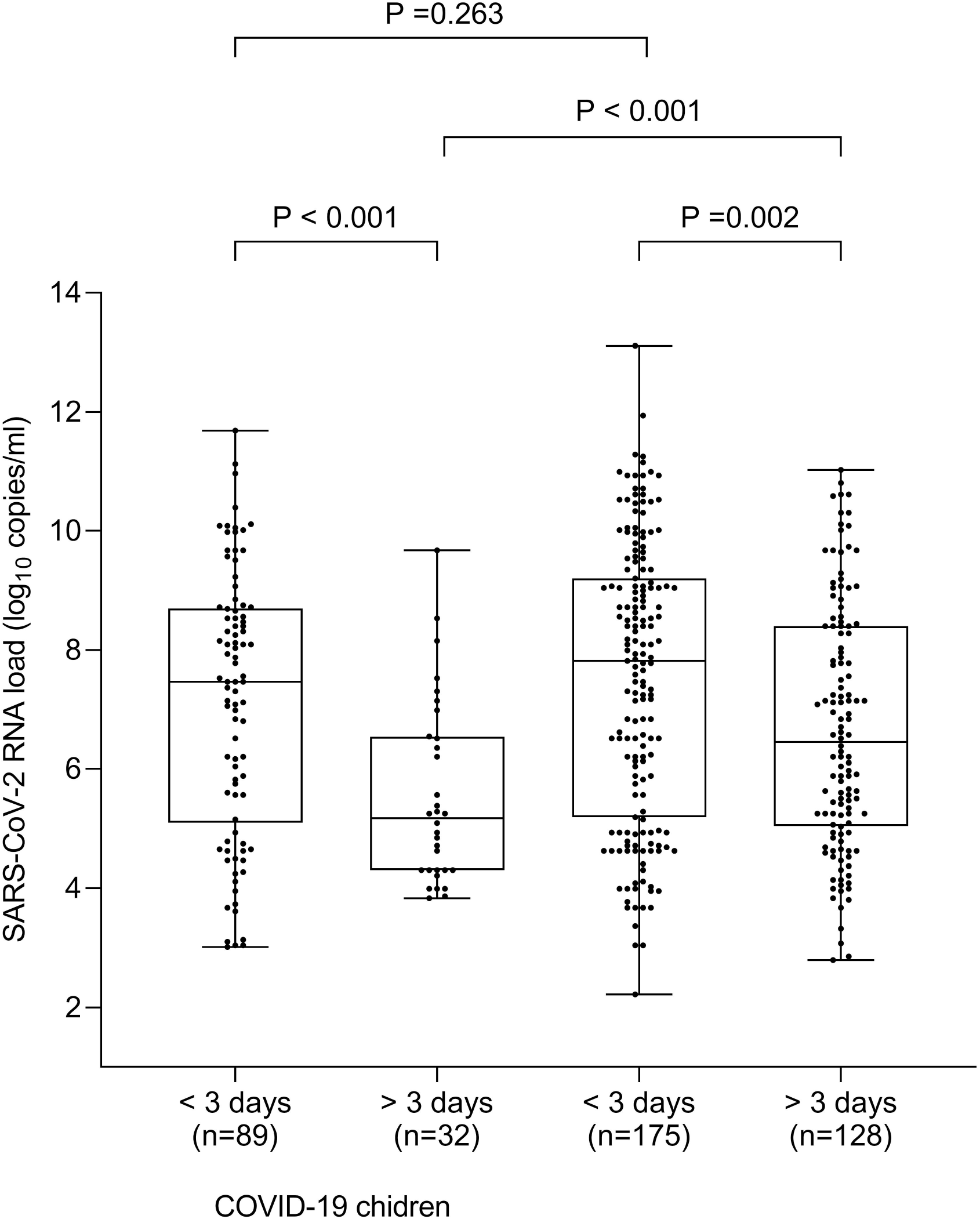
Estimated initial SARS-CoV-2 RNA loads in nasopharyngeal specimens from children and adults with COVID-19 according to the time of sampling after symptoms onset. Medians are indicated by midlines, the top and bottom edges of boxes represent the interquartile range (IQR). Whiskers indicate the upper and lower values. The number of patients in each group as well as *P* values for comparisons between groups (median SARS-CoV-2 RNA levels) are shown.

Finally, we compared initial SARS-CoV-2 RNA loads across age groups conventionally defined for children (infants, toddlers, preschoolers, school-aged, and adolescents) and arbitrarily set for adults (18 to 65 years/>65 years). Pairwise comparison analyses are shown in Fig. 1B. Overall, there were no between-group differences in either children or adults (*P*=>0.14 for all pairwise comparisons). Moreover, as shown in Fig. 3A and 3B, no correlation was found between SARS-CoV-2 RNA loads and patient age, either for children (Rho, 0.008 *P*=0.93) or adults (Rho, 0.005; *P*=0.92).

**Figure 3.**
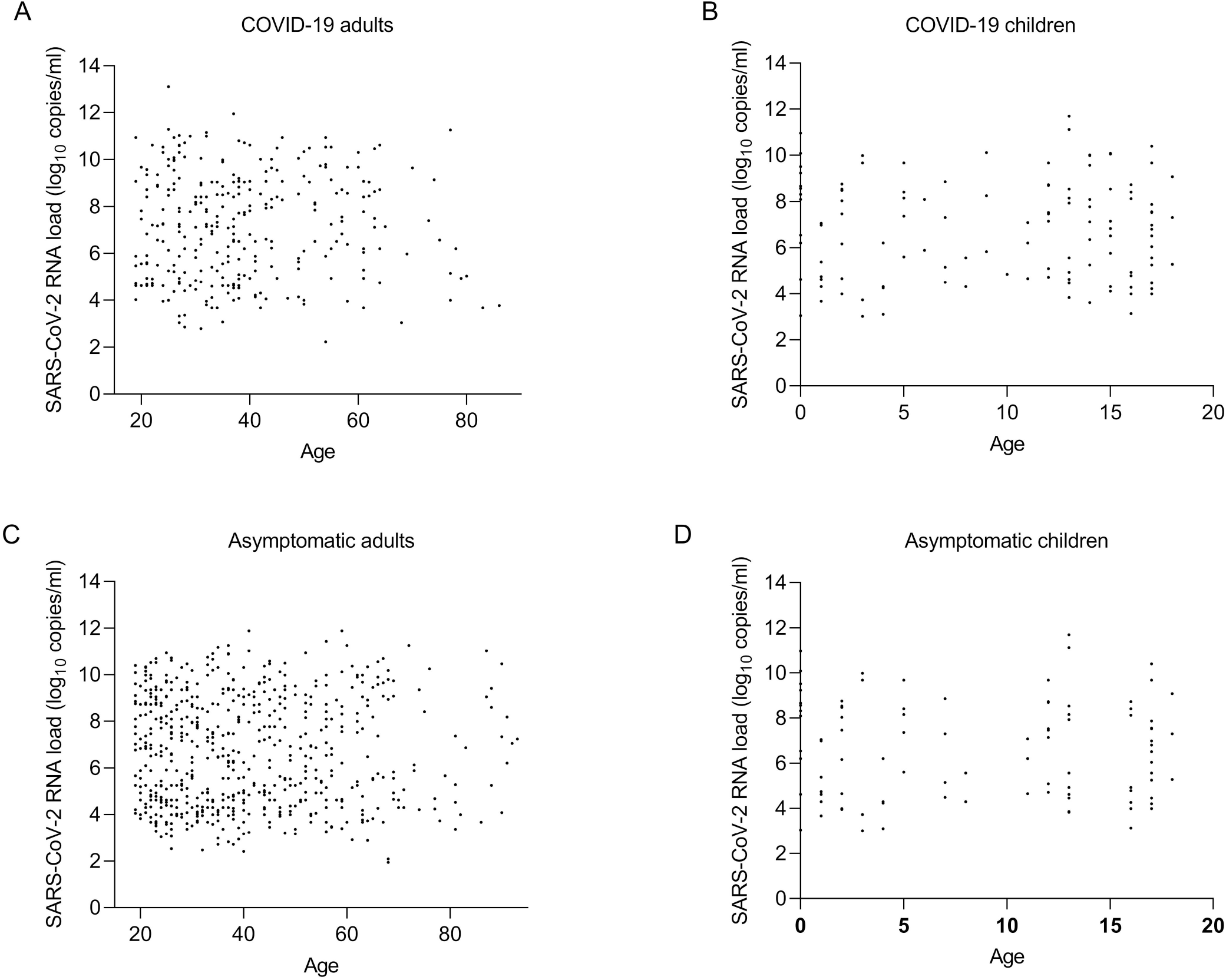
Correlation between estimated initial SARS-CoV-2 RNA load in nasopharyngeal specimens from adults (A) and children (B) with COVID-19, and from asymptomatic adults (C) and children (D) and age of participants.

### SARS-CoV-2 RNA load in asymptomatic children and adults

A wide range of SARS-CoV-2 RNA loads were detected in asymptomatic children and adults (Fig. 4A), likely reflecting the broad spectrum of NP collection times after exposure to the presumed index case which, it should be noted, was not dissimilar between children and adults. SARS-CoV-2 RNA loads in asymptomatic children (median, 6.20 log_10_ copies/ml) and adults (median, 6.48 log_10_ copies/ml) were comparable in magnitude (*P*=0.97). Likewise, no differences in SARS-CoV-2 RNA loads were observed across pediatric ages or between adults aged ≤65 years or older (Fig. 4B) and no correlation was found between age and SARS-CoV-2 load (Rho, 0.066; *P*=0.44 for children and Rho, 0.020; *P*=0.62 for adults (Fig. 3C and 3D).

**Figure 4.**
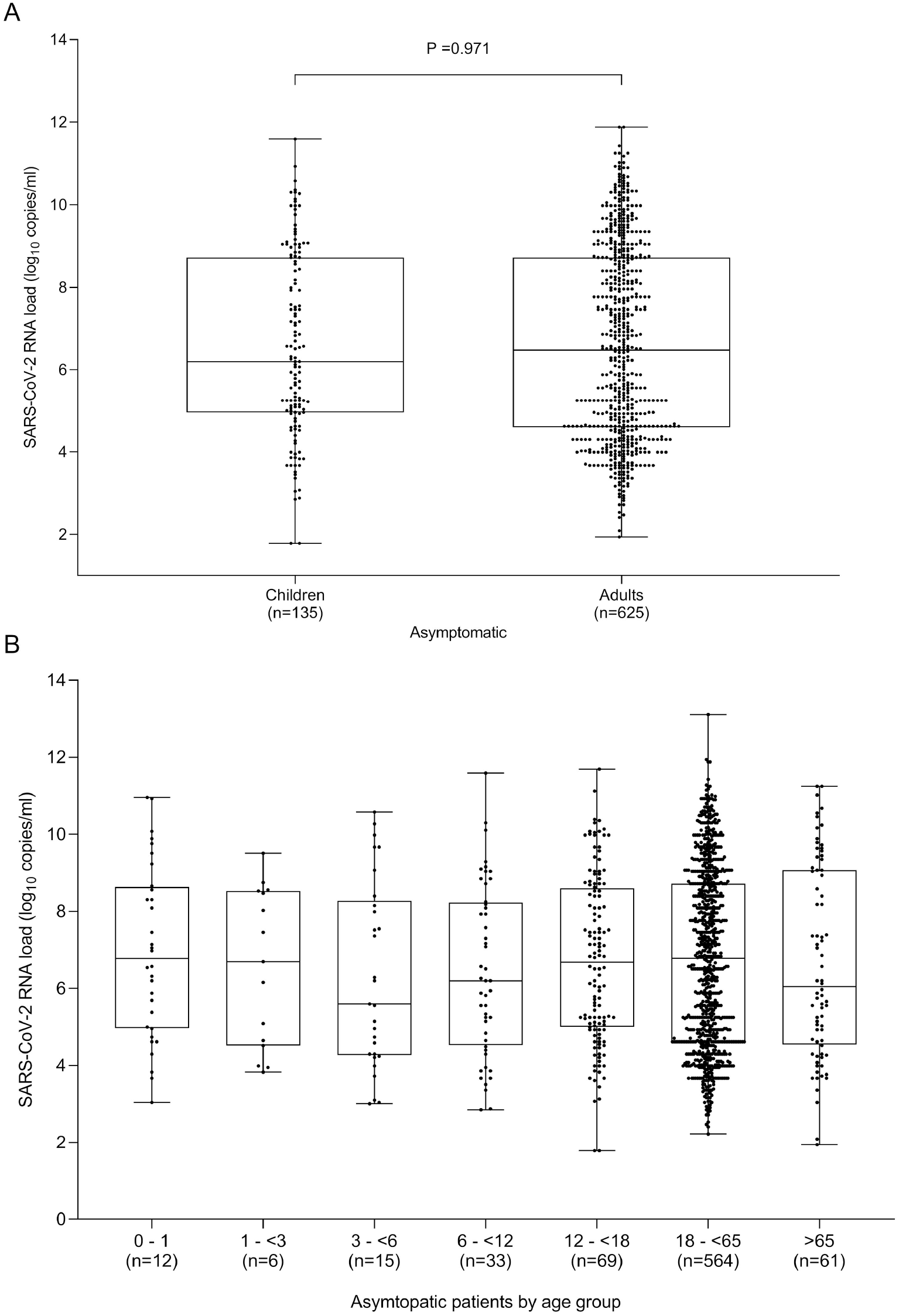
Overall estimated initial SARS-CoV-2 RNA loads in nasopharyngeal specimens from asymptomatic children and adults with COVID-19 (A) and those found across different pediatric and adult ages (B). Medians are indicated by midlines, the top and bottom edges of boxes represent the interquartile range (IQR). Whiskers indicate the upper and lower values. The number of patients in each group as well as *P* values for comparisons between groups (median SARS-CoV-2 RNA levels) are shown.

### Comparison of URT SARS-CoV-2 RNA load in symptomatic vs. asymptomatic children and adults

Children with COVID-19 symptoms displayed slightly higher SARS-CoV-2 RNA load than their asymptomatic counterparts (Fig. 5A), although statistical significance was not reached (*P*=0.61). In adults, median estimated SARS-CoV-2 RNA load was significantly higher in symptomatic than asymptomatic subjects (*P*=<0.001) (Fig. 5B), nevertheless, it was comparable (*P*=0.61) when patients sampled within 48 h after symptoms onset were excluded from the analysis (Supplementary Fig. 1).

**Figure 5.**
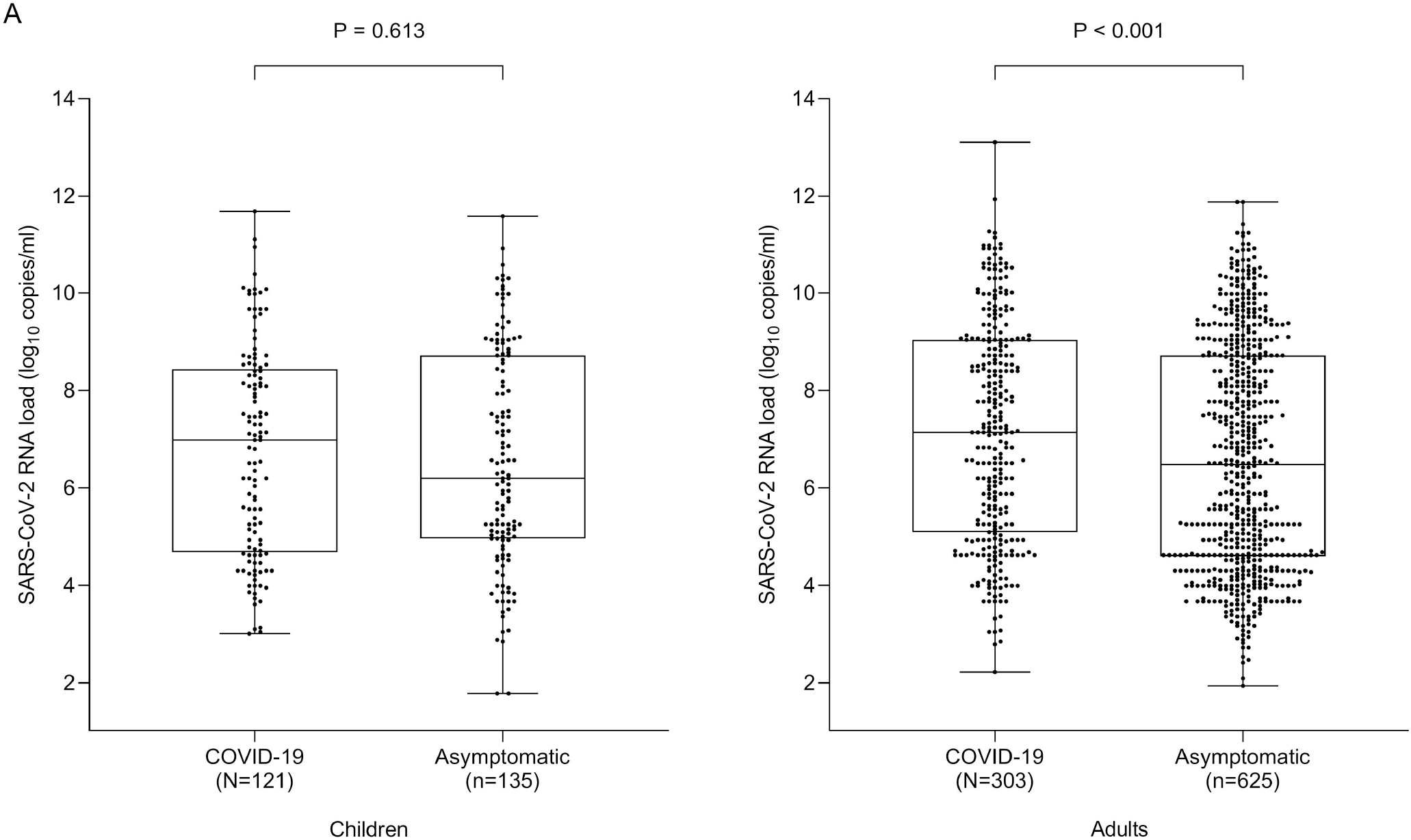
Comparison of estimated initial SARS-CoV-2 RNA loads in nasopharyngeal specimens from children (A) and adults (B) either asymptomatic or presenting with COVID-19. Medians are indicated by midlines, the top and bottom edges of boxes represent the interquartile range (IQR). Whiskers indicate the upper and lower values. The number of patients in each group as well as *P* values for comparisons between groups (median SARS-CoV-2 RNA levels) are shown.

### Inference of the percentage of children and adults presumably shedding infectious virions

We previously reported that SARS-CoV-2 could not be cultured from NP specimens returning C_T_>25 (<5.9 log_10_ copies/ml) by the TaqPath COVID-19 RT-PCR [16]. We investigated the distribution of specimens yielding C_T_ <25 across children and adults. The data are shown in Supplementary Fig. 2. Overall, the percentage of NP specimens returning SARS-CoV-2 N RT-PCR C_T_s below the abovementioned threshold was similar for symptomatic children and adults (P=0.28) and was also comparable between asymptomatic children and adults *(P*=0.87). Among children, that percentage appeared higher for those aged under 3 years. For most age groups the percentage was higher in symptomatic than in asymptomatic subjects, although these differences did not reach statistical significance.

### Assessment of the cellularity of NP specimens collected from pediatric and adult participants

To assess the quality of NP specimens collected from children and adults regarding cellularity, we randomly selected 30 samples from each population group (n=60) that were matched in SARS-CoV-2 RNA load (median 5.70 log_10_ copies/ml; range 3.5-11.6 log10 copies/mL in specimens from children; median, 6.60 log_10_ copies/ml; range, 2.2-10.9 log_10_ copies/ml in specimens from adults; *P*=0.99). These specimens were assayed with an in-house designed RT-PCR amplifying the housekeeping GUSB gene. The C_T_ of NP samples obtained from children and adults did not differ significantly (median C_T_, 28.1; range, 24.8-32.7; and median C_T_, 29.0; range, 25.2-31.7, respectively, *P*=0.3), suggesting that SARS-CoV-2 RNA loads measured in the two population groups were not biased by differences in cellularity across NP specimens.

## DISCUSSION

To our knowledge, this is one of the largest studies to date investigating how children and adults (either asymptomatic or presenting with mild COVID-19 at time of sampling) compare regarding URT SARS-CoV-2 RNA shedding. Several major findings arose from the current study. First, overall, there was no significant difference in initial URT SARS-CoV-2 RNA load between COVID-19 pediatric and adult patients. Furthermore, the percentage of NP specimens potentially yielding infectious virions (C_T_<25), as previously estimated [16], was similar across children and adults. Interestingly, a subanalysis categorizing patients by time to specimen collection since symptoms onset revealed that SARS-CoV-2 RNA loads in children and adults were comparable at early times (within 48 h), when peak levels are known to be reached [19], but were significantly lower in children at later times, suggesting a faster URT SARS-CoV-2 RNA clearance rate in children. In accordance with our data, Baggio et al [10] found similar estimated SARS-CoV-2 loads in children and adults sampled within the first 5 days after onset of symptoms. Likewise, Heald-Sargent et al. [9] found that preschool-and school-aged children sampled within one week after symptoms onset display similar SARS-CoV-2 RNA loads to their adult counterparts. In contrast, a slightly lower SARS-CoV-2 RNA load in children than adults was reported in a German study [8]; however, information on symptom onset was not provided [8].

Second, pairwise comparison analyses revealed no significant differences in SARS-CoV-2 RNA load across age groups, in either symptomatic children or adults; in children, a similar conclusion can be derived from the study by Kociolek and colleagues [12]. In contrast, age-related differences in SARS-CoV-2 RNA load have been reported previously in children [9,10]; specifically, young children (<5 years old) had significantly lower median SARS-CoV-2 RT-PCR C_T_ values than older children and adults. Further studies involving larger cohorts are warranted to explain this apparent discrepancy.

Third, SARS-CoV-2 transmission to susceptible individuals from asymptomatic infected adults has been documented and postulated to facilitate virus dissemination in the community [20–22]. Here, we found no difference either in SARS-CoV-2 RNA loads or the percentage of NP specimens presumably yielding infectious virus between asymptomatic adults and children, irrespective of the age group considered, suggesting that asymptomatic children may contribute to virus spreading to the same extent as adults seemingly do.

Fourth, previous studies reported an overlapping initial SARS-CoV-2 RNA load distribution in symptomatic and asymptomatic adults, regardless of age and baseline medical condition [23,24]. Here, contrarily, we observed higher viral loads in symptomatic than in asymptomatic adults; however, this difference disappeared when excluding patients sampled very early after onset of symptoms from the analyses. While the kinetics of SARS-CoV-2 RNA load in URT has been clearly established in symptomatic individuals, with viral load peaking within 48 h after symptoms onset, it remains to be precisely characterized in asymptomatic subjects; as a result, between-group differences in results in the latter subset likely depend upon the time window of specimen collection. Regarding children, we found similar SARS-CoV-2 loads in symptomatic and asymptomatic individuals, although a subtle trend towards higher viral loads was seen in the former. Our data concur with those of Hurst and et al.^25^, but are in contradiction to those of Kociolek et al.^12^ which clearly pointed to lower SARS-CoV-2 RNA loads in asymptomatic children than in those with mild to moderate COVID-19. In this regard, it must be stressed that in our study asymptomatic individuals were tested relatively soon after exposure, whereas in Kociolek’s the authors admit a potential population bias towards lower SARS-CoV-2 loads due to an excessive number of remote infections detected via screening programs (i.e. hospital pre-admission).

Like the majority of commercially-available SARS-CoV-2 RT-PCRs, the RT-PCR assays used in the current study do not co-amplify a housekeeping gene, thus precluding assessment of sample cellularity. Given the widely varying quality of NP specimens [26] which impacts significantly on estimated SARS-VoV-2 RNA loads [26], we compared a randomly selected set of NP specimens from children and adults for their cellular content using a housekeeping-gene RT-PCR set in parallel. We found overlapping CTs in samples from both subject groups, making it unlikely that differences in cellularity had a major impact on our results. However, only a small number of NP specimens were screened for their cellular content.

The current study has several limitations. First, clinical outcome of asymptomatic individuals, which may be determined by peak viral load, could not be ascertained in a large number of participants. Second, only initial SARS-CoV-2 loads were taken into consideration in the analyses, so that we could not have captured the true virus replication rate on an individual basis. Third, no attempt was made to subcategorize individuals according to their baseline medical condition.

In summary, we conclude that SARS-CoV-2 RNA loads in non-hospitalized or asymptomatic COVID-19 children of all ages were comparable to those estimated in adults. Our findings indicate that children may spread SARS-CoV-2 in the general population at the same level as adults.

## Data Availability

The data that support the findings of this study are available on request from the corresponding author, DN.

## ACKNOWLEDGMENTS

We thank all personnel working at Health Department Clínico-Malvarrosa for their unwavering commitment in the fight against COVID-19. Eliseo Albert holds a Río Hortega research contract from the Carlos III Health Institute (Ref. CM18/00221).

## AUTHOR CONTRIBUTIONS

RC, FB, EA, IT, DS, CP and JC: Methodology and data collection. RC, FB: Formal analysis. RC, FB, CM-C and DN: Conceptualization and validation of data. S C-S, AB-F, MILC, JRB-M and CM-C were phsycians in charge of children. DN: writing the original draft. All authors reviewed and approved the original draft.

## FIGURE LEGENDS

**Supplementary Figure 1**. Comparison of estimated initial SARS-CoV-2 RNA loads in nasopharyngeal specimens from adults either asymptomatic or presenting with COVID-19 within 48 h after onset of symptoms. Medians are indicated by midlines, the top and bottom edges of boxes represent the interquartile range (IQR). Whiskers indicate the upper and lower values. The number of patients in each group as well as *P* values for comparisons between groups (median SARS-CoV-2 RNA levels) are shown.

**Supplementary Figure 2**. Percentage of nasopharyngeal specimens from children or adults across different age groups returning RT-PCR CTs <25.

