## Supplementary figures and images for "Upper respiratory tract SARS-CoV-2 RNA loads in symptomatic and asymptomatic children and adults"

### Supplementary Table 1

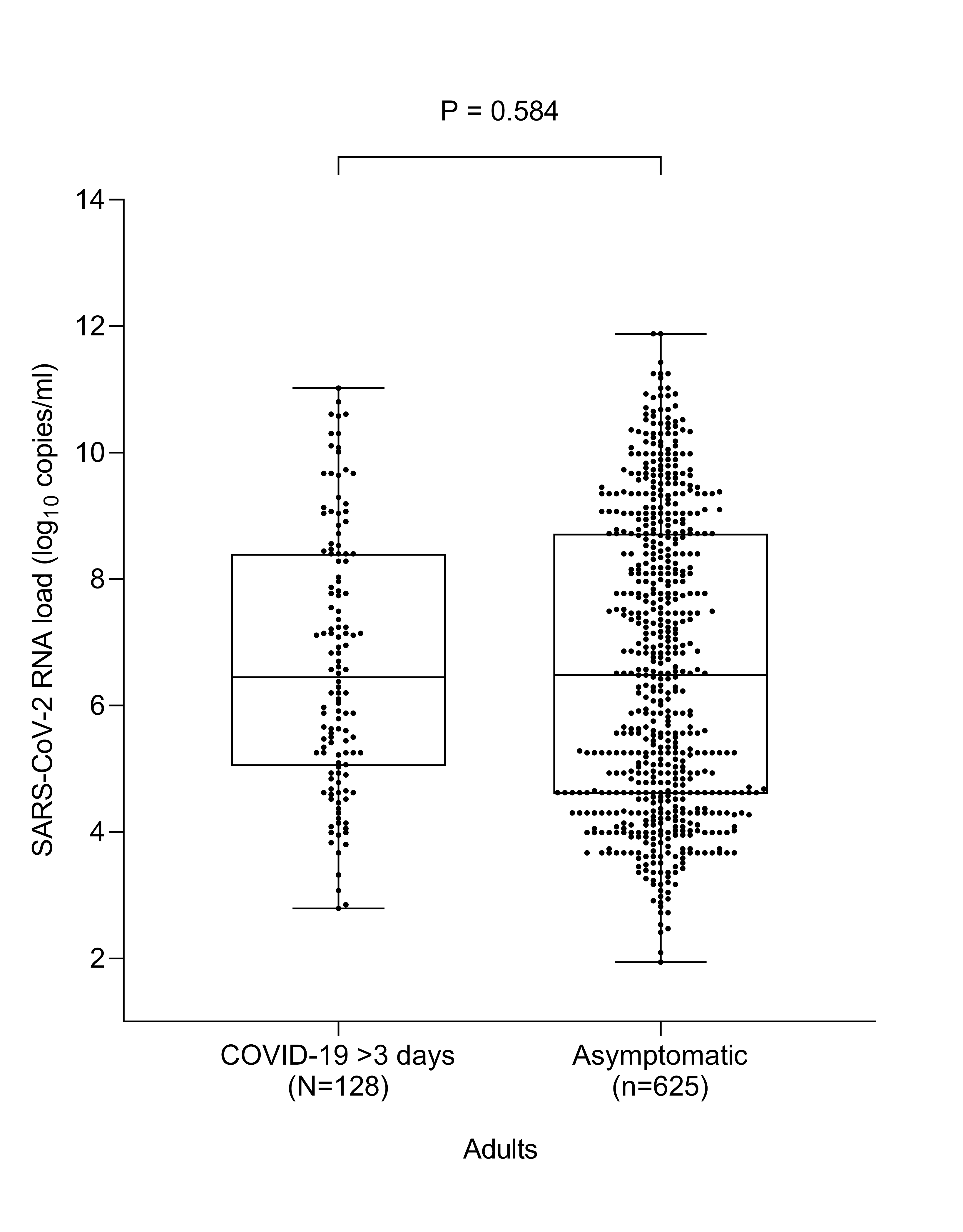

### Supplementary Table 2

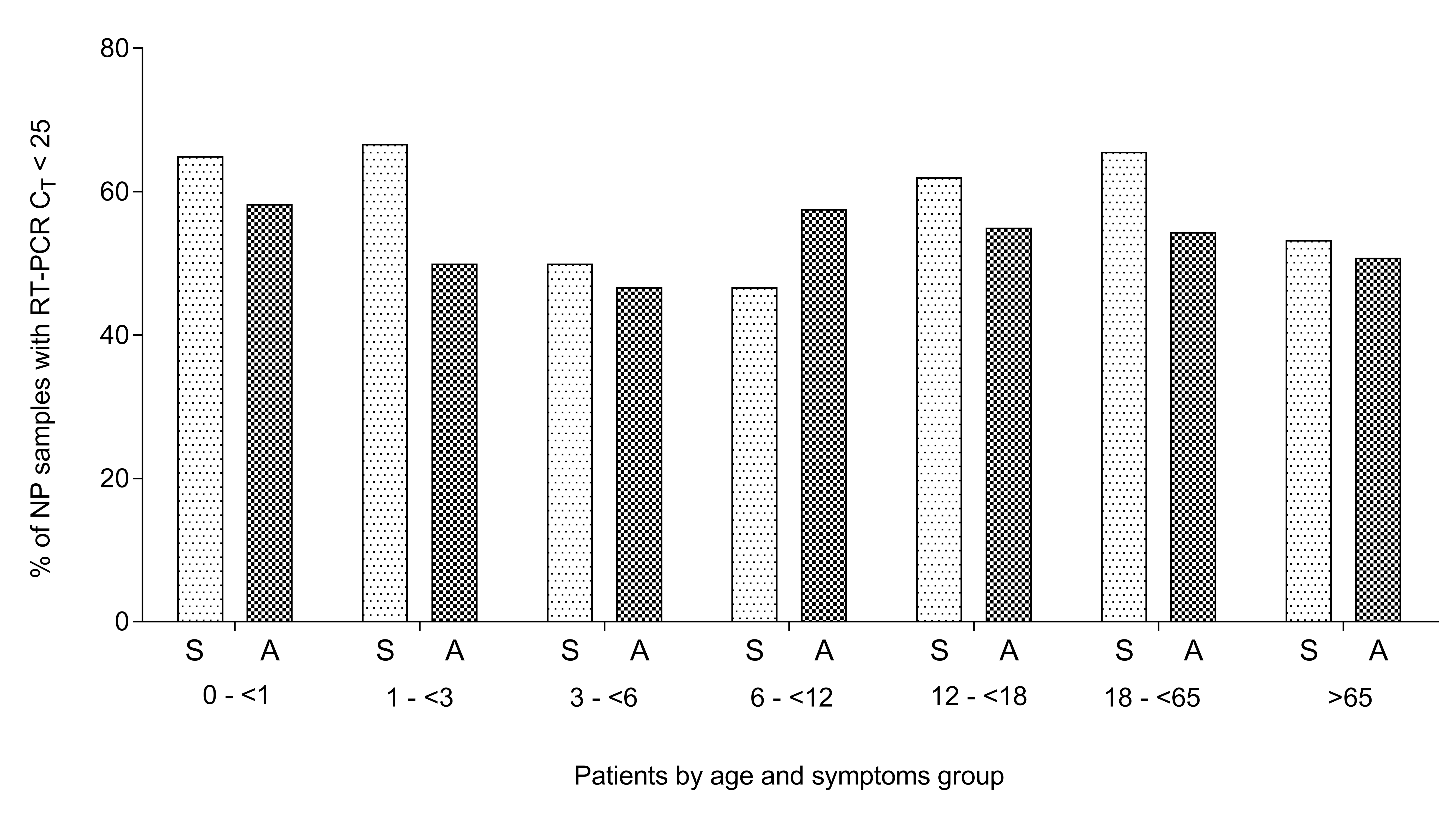
